# Who is dying from Covid-19 in the United Kingdom? A review of cremation authorisations from a single South Wales’ crematorium

**DOI:** 10.1101/2020.07.01.20136317

**Authors:** Roland Salmon, Stephen Monaghan

## Abstract

**Background:** Covid 19 is pandemic in the UK. To date only studies in the UK on hospital deaths have been published in the peer reviewed literature. Legal requirements for cremation in England and Wales require the collection of information that can be used to improve understanding of Covid 19 deaths in both hospital and community settings.

**Aim:** To document demographic and clinical characteristics, including likely place of infection, of individuals dying of Covid 19 to inform public health policy

**Design:** A comprehensive case series of deaths from Covid 19 between 6 April and 30 May.

**Setting:** A crematorium in South Wales

**Participants:** Individuals for whom an application was made for cremation.

**Main outcome measures:** Age, sex, date and place of death, occupation, comorbidities, where infection acquired.

**Results:** Of 752 cremations, 215(28.6%) were Covid-19 of which 115 (53.5%) were male and 100 (46.5%) female. The median age was 82 years, with the youngest patient being 47 years and the oldest 103 years. Over half the deaths (121/215: 56.3%) were over 80 years. Males odds of dying in hospital, rather than the community were 1.96 times that of females (95% Confidence Intervals (CI) 1.03 −3.74, p=0.054) despite being of similar age and having a similar number of comorbidities. Only 21(9.8%) of 215 patients had no comorbidities recorded. Patients dying in nursing homes were significantly older than those dying in hospital(median 88y (IQ range 82-93y) v 80y (IQ range 71-87y): p<0.0001). Patients dying in hospital had significantly more comorbidities than those dying in nursing homes (median 2: IQ range 1-3 v. 1: IQ range 1-2: p <0.001).

**Conclusions:** In a representative series, comprising both hospital and community deaths, persons over 80 with an average 2 comorbidities predominated. Although men and women were represented in similar proportions, men were more likely to die in hospital. Over half the infections were acquired in either hospitals or nursing and residential homes with implications for the management of the pandemic, historically and in the future.

## Background

Covid-19, a viral infectious disease caused by the SARS CoV 2 virus emerged in China at the end of 2019 and was declared pandemic by the World Health Organisation on 11 March 2020^1^. The first case occurred in the UK on 31st January 2020 and in Wales, where 1,441 deaths have occurred as of 12 June 2020^2^, on 28th February 2020^3^. The disease has given rise to a number of far-reaching public health measures. So far, in the UK, there have been three large community studies, of which in two, the main outcome is deaths in hospital^4,5^ and in the third positive SARS CoV 2 tests^6^. There has also been a small study of risk factors for intensive care unit admissions in South Wales^7^. All identify similar risks for mortality and morbidity; increasing age, male gender, diabetes, chronic heart, lung, kidney or neurological disease, malignancy and dementia. Importantly, although there has been an ecological study comparing death rates, including out-of-hospital deaths, by local authority area, in Great Britain^8^, no UK studies, to date, have reported the characteristics and risks of death in individuals dying from COVID-19 that includes those patients dying out-of-hospital, in nursing and residential homes or at home, as well as those dying in hospital.

In 2012, as a result of The Cremation (England and Wales) Regulations 2008^9^, themselves arising out of the Inquiry into Harold Shipman^10^, a new suite of forms for the authorisation of cremation came into use. Although procedures have been temporarily simplified to assist the management of the Covid-19 pandemic^11^, the key information requirements are unchanged. These emphasise that the fact and cause of death has been definitely ascertained and require a brief text account (question 9, form 4) of the “symptoms and other conditions” that led to the conclusions about the cause of deaths. This account, although unstructured, permits a view of the course of the final illness which can be used better to understand deaths from Covid-19.

Cardiff Thornhill Crematorium is a Local Authority run crematorium (run by Cardiff Council, one of 22 unitary local authorities in Wales) that performed 2,850 cremations in 2017, a typical year in terms of activity.

## Objectives

To characterise those individuals dying from Covid-19, that were authorised for cremation, in terms of age, sex, occupation, comorbidities and where the infection had been acquired, in order better to inform public health policy to address the infection.

## Methods

Information was taken from: Cremation Form 4 in which a certifying clinician is required both to record the cause of death (question 11) in a format that reflects the separate Medical Certificate of Cause of Death (MCCD) and to give a brief account (question 9) of the “symptoms and other conditions” that led them to that conclusion: Cremation Form 1 in which the applicant for cremation (usually the next of kin) gives the age and occupation of the deceased. Both forms record the home address, date and time of death. Ethnicity, however, is not recorded.

Deaths were defined as Covid-19 if specifically mentioned in response to q.11 or if a SARS CoV2 positive test was documented in response to q.9. Comorbidities were any relevant conditions mentioned in answer to either of questions 9 or 11.

The authors authorised all patients over the study period. Data was entered onto a structured *pro forma;* sex, age, occupation, date of death, location of death (hospital, institutional nursing or residential home, own home), comorbidities and whether infection was acquired in hospital (defined as occurring 6 days after admission OR following admission for an unrelated condition). Nursing and residential home residents were assumed to have acquired their infection in their nursing or residential home unless they fulfilled the criteria for having acquired the infection in hospital.

Analysis was performed in EpiInfo version 7(US Centers for Disease Control and Prevention (CDC))^12^. Median and interquartile ranges were calculated for age and the number of comorbidities; simple frequencies for other variables. Differences in patient characteristics and comorbidities (i) between men and women and (ii) by place of death were examined using the Mann-Whitney U test for continuous variables and the chi squared test (Yates corrected) for categorical variables.

Patients and the public were not involved in this study, responding, in a timely way, to a Public Health Emergency of International Concern.

## Results

Of the total of 752 cremations authorised over the period of 6th April until 29th May, 215(28.6%) were Covid-19 of which 115 (53.5%) were male and 100 (46.5%) female (p=NS). Dates of death are shown in Figure 1. The median age was 82 years, with the youngest patient being 47 years and the oldest 103 years. Over half the deaths (121/215: 56.3%) were over 80 years. Among 146 patients who died in hospital, 85 (58.2%) were male and 61 (41.8%) were female (p NS). Males’ odds of dying in hospital, rather than the community were 1.96 times that of females (95% Confidence Intervals (CI) 1.03 −3.74, p=0.054) despite being of similar age (males: median age 81y: interquartile (IQ) range 72y-87y v females: 84y: IQ range 72-90.5y) and having a similar number of comorbidities (both sexes: median 2, range 0-7). Only 21(9.8%) of 215 patients had no comorbidities recorded.

**Figure 1.**
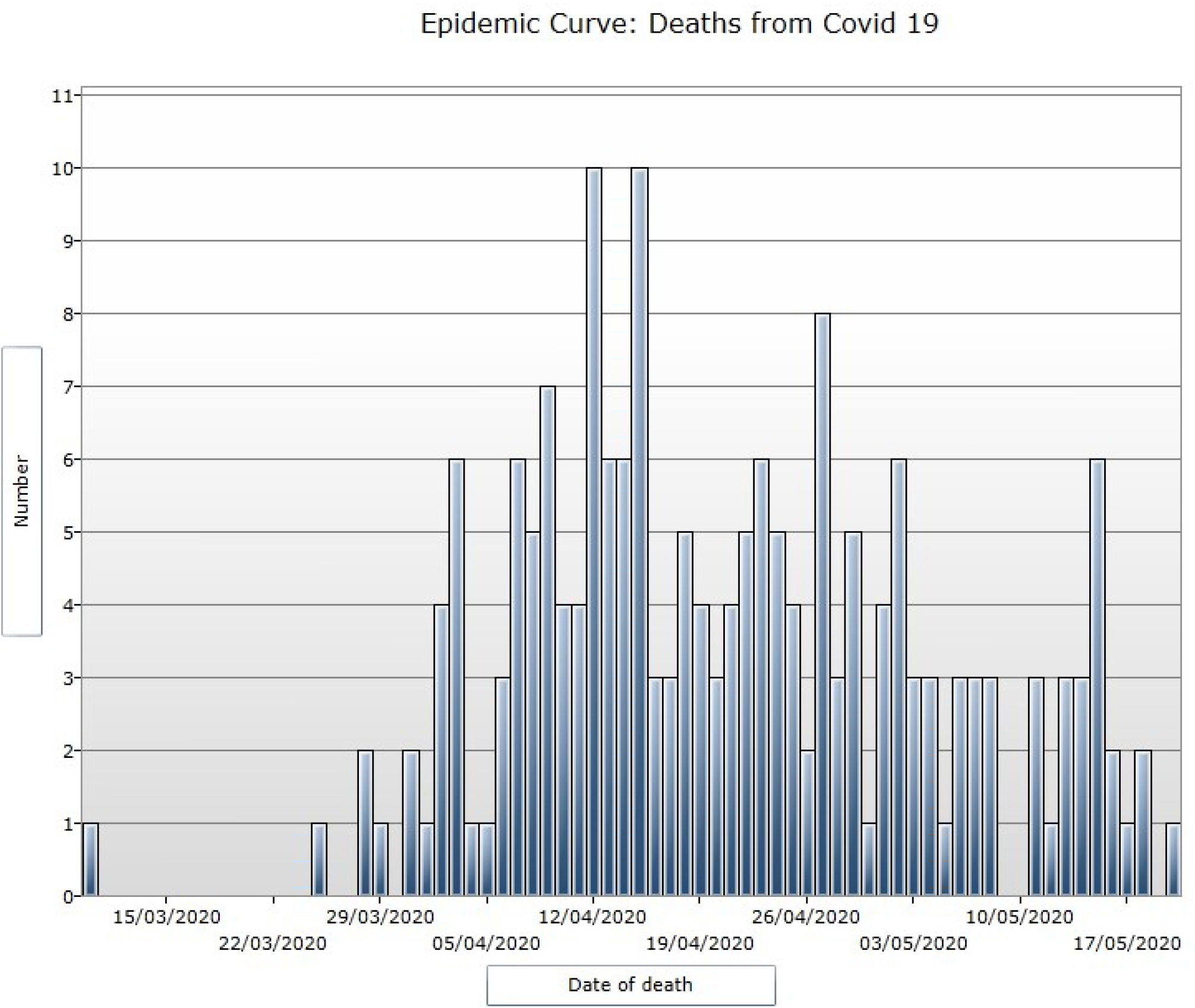

Patients’ comorbidities, by place of death, are shown in Table 1. Patients dying in nursing homes were significantly older than those dying in hospital(median 88y (IQ range 82-93y) v 80y (IQ range 71-87y): p<0.0001). Patients dying in hospital had significantly more comorbidities than those dying in nursing homes (median 2: IQ range 1-3 v. 1: IQ range 1-2: p <0.001). Patients dying in hospital were significantly more likely than those dying in nursing homes, to have chronic heart disease (p<0.05) or chronic pulmonary disease, excluding asthma (p<0.05) but significantly less likely to have dementia (p<0.00005). No patient dying in their own home had dementia (p<0.001).

**Table 1.**
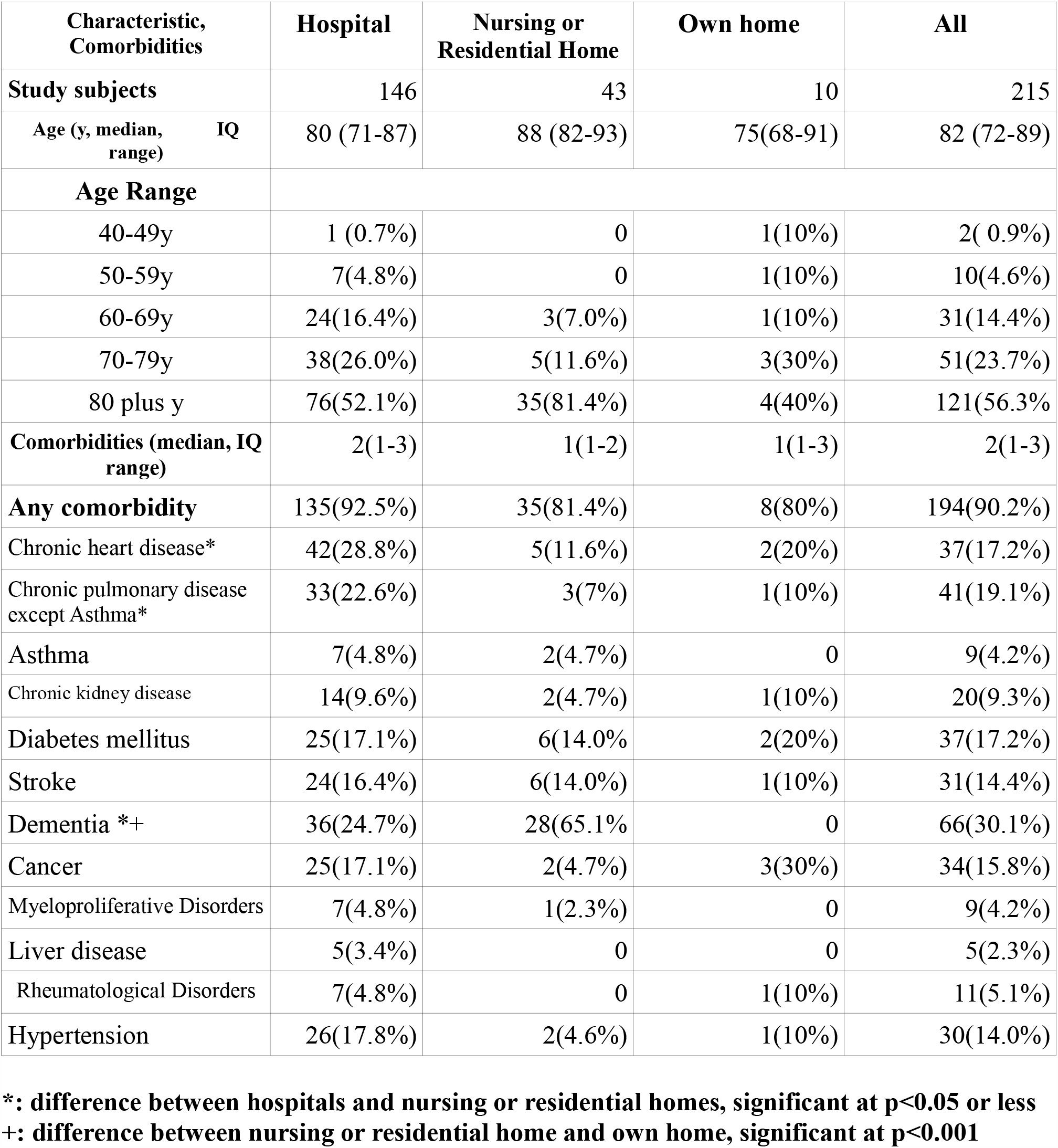
Characteristics and comorbidities in patients dying of Covid-19, by place of death.

Of the 31(14%) patients less than 65y, 20 were recorded as working and occupations included 2 NHS, 2 care sector and 1 transport sector staff.

Of 215 cases of Covid-19, 63 (29.3%) of infections were hospital acquired and a further 55(25.6%) acquired in nursing or residential homes.

## Discussion and Conclusions

This is the first comprehensive study of deaths, including individuals dying both in hospital and in the community, in the United Kingdom. Other studies have either looked at patients dying in hospital^4,5,7^ or patients testing positive for SARS CoV 2 in the community^6^ or have been ecological studies^8^.

The study uses the information required by a crematorium, under the law of England and Wales^9^, before a person can be cremated, information which includes a brief clinical account to support the stated causes of death. Although unstructured text, official guidance on completion of the forms does exist^13^ and this information can give a rounded picture of the circumstances leading to a death, similar to a clinical referral letter. Cremation forms, in this way, represent a rich source of information on the end of life and elements such as the type and appropriateness of care. This time-honoured and legally laid down process, can thus be used to promote health and prevent disease. To utilise better, this would require a degree of central organisation at regional or national level. The introduction of the Medical Examiner system, in England and Wales^14^ may present an opportunity to do this but hitherto the focus has been almost entirely on patient satisfaction (or, more strictly, that of their relatives) and healthcare quality. Whilst important, this ignores the usages that mortality statistics have been put to, historically, to tackle other areas of public health such as health protection and health promotion, such as Clean Air Acts, following the 1953 London smog or the current tracking of Covid-19. In fact, better collation of mortality statistics and more extensive and systematic recording of clinical, pathological and risk factor data and linking those mortality statistics with other public data sources (*eg* cancer registries, prescribing data, hospital episode statistics, air quality data) would allow the contemporary quantification of several “big ticket” current public health issues, other than Covid-19, such as, alcohol use, obesity, anti-microbial resistance and air pollution.

The denominator population, from which patients that use Cardiff Thornhill crematorium are drawn, is difficult to characterise exactly, as more than one crematorium serves the Cardiff Council area and equally cremations are accepted from other local authority areas. Cardiff itself has a population of 335,145 persons, according to the 2011 census and Thornhill crematorium users are thought broadly to reflect this.

Deaths occurred, as in other studies, worldwide, mainly in the elderly (median age 82y) with over half in those over 80 years old and with over 90% having pre-existing medical conditions. The proportion of men and women in the whole group did not differ significantly, unlike the other UK studies, restricted to deaths in hospital. However, an excess of male deaths, in the subset of deaths occurring in hospital, was similar to these other UK studies. The odds of men dying in hospital was thus nearly twice that of women, even though they were of similar age with a similar number of comorbidities. This phenomenon is unexplained and it will be of interest to know if it is replicated in other settings where deaths in the community are also recorded.

Over half the infections, plausibly, were acquired in institutional care settings, of which nearly 30% were acquired in hospital. Concerns have been raised about hospital transmission in other UK settings but in the absence of official figures, the proportion has been estimated as 5-20%^15^. This inevitably poses the question of what proportion of these could have been prevented by more effective infection control procedures. It is particularly sobering to compare these percentages with the purported benefits of Non-Pharmaceutical Interventions (NPIs), as modelled by Imperial College, London,^16^ which were so influential in guiding UK government policy at the start of the pandemic. The biggest predicted percentage reduction in deaths, for any of the combinations of NPIs was also 50%. The implication of this is that more focussed interventions to prevent the introduction of SARS CoV 2 and to control its spread, in healthcare settings, may have had the same potential to reduce deaths as the general social distancing and other costly measures that were, in the event, introduced. It poses the question whether the UK’s high mortality, when compared with other European countries^17^, has as much to do with its historically high hospital occupancy rates^18^ and underfunding of the social care sector^19^ as it does with the, widely blamed, delay in introducing lockdown^20^. Indeed, given the apparent ineffectiveness of the NPIs in keeping mortality down, a focus on control of SARS CoV 2 infection in hospitals, nursing and residential homes might have been a more effective approach in limiting deaths and certainly would not have resulted in the same social and economic disruption as lockdown and general social distancing. Going forward, this may still represent the most effective use of testing and tracing teams rather than attempting to extinguish spread in a wider community of people who would, largely, be expected to recover from the infection without mishap.

## Conclusion

Cremation certificates represent a useful source of information on health problems locally and could, with greater co-ordination, contribute to a national picture. In a representative series of deaths, in persons, authorised for cremation, in South Wales, comprising both hospital and community deaths, persons over 80 with an average 2 comorbidities predominated. Although, unlike most other studies, there were similar proportions of men and women overall, men were more likely to die in hospital. Over half the infections were acquired in either hospitals or nursing and residential homes with implications for the management of the pandemic, historically and in the future.

## Data Availability

The data set would, ordinarily, be shared on application to the authors.

## Acknowledgements

Bereavement Services staff, Cardiff Council.

Sian King, Evidence Service, Public Health Wales, Swansea

## Data sharing

The data set would, ordinarily, be shared on application to the authors.

## Transparency Statement

Dr Salmon, the lead author (the manuscript’s guarantor) affirms that the manuscript is an honest, accurate, and transparent account of the study being reported; that no important aspects of the study have been omitted; and that any discrepancies from the study as planned (and, if relevant, registered) have been explained.

## Ethical approval

This study uses information required under the Cremation (England and Wales) Regulations 2008, which may be used for the purposes of the public interest.

## Contributions

Dr Salmon conceived the study, collected and analysed the data and edited the text. Dr Monaghan advised on study design, collected data and edited the text.

## Appendix 1

STROBE Statement -checklist.

STROBE Statement - checklist
|  | Item No. | Where found (page) |
| --- | --- | --- |
| **Title and abstract** | 1 | 1,2 |
| **Introduction** |  |  |
| Background/Rationale | 2 | 2,3 |
| Objectives | 3 | 3 |
| **Methods** |  |  |
| Study design | 4 | 3 |
| Setting | 5 | 3 |
| Participants | 6 | 3 |
| Variables | 7 | 3 |
| Data sources/measurement | 8 | 3 |
| Bias | 9 | 3 |
| Study size | 10 | 3 |
| Quantitative variables | 11 | 3 |
| Statistical methods | 12 | 3 |
| **Results** |  |  |
| Participants | 13 | 3 |
| Descriptive data | 14 | 3,4, Table 1, Figure 1 |
| Outcome data | 15 | 3,4, Table 1 |
| Main results | 16 | 3,4, Table 1 |
| Other analyses | 17 | None |
| **Discussion** |  |  |
| Key results | 18 | 4,5 |
| Limitations | 19 | 4,5 |
| Interpretation | 20 | 4,5 |
| Generalisability | 21 | 4,5 |
| **Other information** |  |  |
| Funding | 22 | 6 |

